# Subtyping of Type 2 Diabetes from a large Middle Eastern Biobank: Implications for Precision Medicine

**DOI:** 10.1101/2024.12.26.24319530

**Authors:** Nayra M. Al-Thani, Shaza B Zaghlool, Abdul Badi Abou-Samra, Karsten Suhre, Omar M E Albagha

## Abstract

**Background:** Type 2 diabetes (T2D) can be classified into Severe Insulin-Deficient Diabetes (SIDD), Severe Insulin-Resistant Diabetes (SIRD), Mild Obesity-related Diabetes (MOD), and Mild Age-related Diabetes (MARD). This classification predicts disease complications and determines the best treatment for individuals. However, the classification’s applicability to non-European populations and sensitivity to confounding factors remain unclear.

**Methods:** We applied k-means clustering to a large Middle Eastern biobank cohort (Qatar Biobank; QBB, comprising 13,808 individuals; 2,687 with T2D). We evaluated the efficacy of the European cluster coordinates and analyzed the impact of using actual age on clustering outcomes. We examined sex differences, analyzed insulin treatment frequency, investigated the clustering of maturity-onset diabetes of the young (MODY), and evaluated the incidence of chronic kidney disease (CKD) among T2D subtypes.

**Results:** We identified the four T2D subtypes within a large Arab cohort. Data-derived centers outperformed European coordinates in classifying T2D. The use of actual age, as opposed to age of diagnosis, impacted MOD and MARD classification. Obesity prevalence was significantly higher in females, however, that did not translate to worse disease severity, as indicated by comparable levels of HbA1C and HOMA2-IR. Insulin was predominantly prescribed for individuals in SIDD and SIRD, which also displayed the highest risk of CKD, followed by MOD. Interestingly, most MODY individuals were clustered within MARD, further highlighting the need for precise classification and tailored interventions.

**Conclusion:** The observed sex differences underscore the importance of tailoring treatment plans for females compared to males. For SIDD and SIRD individuals, who are at a higher risk of CKD, insulin therapy requires closer monitoring and physician oversight. Additionally, in populations without access to genetic testing, likely MODY individuals can be identified within the MARD cluster. These findings strongly support the need for a transition to more personalized, data-driven treatment approaches to minimize diabetes-related complications and improve patient outcomes.

## Introduction

The International Diabetes Federation predicts that by 2030, over 600 million individuals will be living with diabetes (1). Type 2 diabetes (T2D) is characterized by dysregulation of beta-cell function and insulin resistance, posing an increased risk for complications such as cardiovascular and renal failure (2, 3). Traditional classification systems do not facilitate individualized treatment for T2D patients. In 2018, a novel scheme proposed categorizing T2D into four subtypes based on six measured variables: Hemoglobin A1C (HbA1C), body mass index (BMI), age of diagnosis, glutamate decarboxylase antibodies (GADA), homeostasis model assessment (HOMA) 2 of estimating beta-cell function (HOMA2-%B), and insulin resistance (HOMA2-IR) (4). This framework, which was developed using the Swedish All New Diabetics in Scania (ANDIS) cohort, has gained traction and offers valuable insights into potential complications associated with T2D (5–10).

The identified subtypes include Severe Autoimmune Diabetes (SAID), Severe Insulin-Deficient Diabetes (SIDD), Severe Insulin-Resistant Diabetes (SIRD), Mild Obesity-related Diabetes (MOD), and Mild Age-Related Diabetes (MARD). Classifying T2D individuals facilitates a precision medicine approach, allowing for tailored interventions to potentially slow the progression of complications.

Recently, we have applied clustering to the Qatar Biobank (QBB), revealing unique proteomic and metabolomic signatures among the identified subtypes (11). Notably, SIDD individuals exhibited autoimmune features linked to complement system activation, while SIRD individuals displayed impaired insulin signaling. MOD individuals showed elevated levels of fatty acid-binding protein and leptin, and individuals classified as MARD exhibited the healthiest profiles.

Numerous studies suggest that subtype clustering is dynamic, that individuals potentially shift between clusters over time and show varying outcomes across different populations (9). For example, research in the Indian population replicated SIDD and MARD subtypes, but they also identified two novel clusters (5). In the Japanese population, individuals classified as SIRD were diagnosed at younger ages and had the highest BMI, a pattern not observed in the MOD subtype (8). Insulin therapy is commonly prescribed for those with severe HbA1C levels, in recent findings, usage of insulin was higher in SIDD, SIRD, and MARD subtypes (8). It has been reported previously that chronic kidney disease (CKD) risks are more prevalent in SIRD (4, 12, 13). Contradicting studies reported SIRD, SIDD, and MOD had a higher prevalence (12), while the other study reported the prevalence is higher in SIDD and MARD (13). The inconsistent results of CKD risk across subtypes suggest that classification can define individuals with a higher risk of CKD in the severe clusters (SIDD and SIRD) but not the mild clusters (MOD and MARD). Moreover, misclassifying individuals with maturity-onset diabetes of the young (MODY) as T2D requires genetic testing for accurate classification, as demonstrated in the QBB cohort (14).

In this study, we applied the Ahlqvist et al. clustering scheme to an extended QBB cohort comprising 13,808 individuals. Our analysis evaluated the applicability of European-derived coordinates, investigated the influence of using actual age versus age at diagnosis on T2D classification, and examined sex-based differences across subtypes within the cohort. Furthermore, we analyzed insulin prescription patterns among T2D subtypes, explored the clustering of MODY individuals, and assessed CKD risk across the QBB cohort.

## Results

### Replication of T2D subtypes in the extended QBB cohort

In this study of the extended QBB cohort, which includes 13,808 participants, 2,765 were identified with T2D (20%) and 71 with T1D (0.5%) as shown in Table 1.

**Table 1:** Comparison of the QBB cohort. The merged data set results in the QBB 13,808 cohort. The data indicate numbers with percentages and means with standard deviation as appropriate. A two-sided t-test is used to determine whether there is a significant difference between the two batches (p values listed in the last column).

| Cohort | QBB 13,808 | p-value |
| --- | --- | --- |
| Sample Size (N) | 13,808 | - |
| Sex (male%) | 6150 (44.5%) | - |
| Age (years) | 41.1 (12.7) | $3.1 \times 10^{-3}$ |
| Age of diagnosis | 38.4 (12.09) | $5.9 \times 10^{-1}$ |
| Type 2 diabetes | 2765 (20.02%) | - |
| T2D Sex (male%) | 1156 (41.8%) | - |
| Type 1 diabetes | 71 (0.5%) | - |
| T1D Sex (male%) | 34 (47%) | - |
| Physician diagnosis | 2467 (17.86%) | - |
| Diabetes treatment | 1714 (12.4%) | - |
| fasting > 8 hrs | 8149 (59.01%) | - |
| Glucose > 200 mg/dL | 508 (3.67%) | - |
| HbA<sub>1c</sub> > 6.5% | 1816 (13.15%) | - |
| BMI (kg/m<sup>2</sup>) | 29.9 (5.9) | $8.9 \times 10^{-3}$ |
| LDL-C (mmol/L) | 3.19 (0.77) | $6.03 \times 10^{-6}$ |
| HDL-C (mmol/L) | 1.39 (0.37) | $1.174 \times 10^{-11}$ |
| Total cholesterol (mmol/L) | 5.18 (0.86) | $1.6 \times 10^{-3}$ |
| Triglyceride (mmol/L) | 1.32 (0.76) | $1.55 \times 10^{-8}$ |
| C-peptide (ng/ml) | 2.3 (1.2) | $7.63 \times 10^{-8}$ |
| C-peptide fasting | 2.16 (1.001) | $4.56 \times 10^{-17}$ |
| Insulin (uU/ml) | 13.6 (16.15) | $2.2 \times 10^{-3}$ |
| Fasting insulin (uU/ml) | 12.4 (28.3) | $4.8 \times 10^{-1}$ |
| Glucose (mmol/L) | 5.4 (2.1) | $4.487 \times 10^{-21}$ |
| Fasting glucose (mmol/L) | 5.6 (2.09) | $1.01 \times 10^{-13}$ |
| HbA<sub>1c</sub> (%) | 5.76 (1.208) | $6.3 \times 10^{-1}$ |
| HOMA2-%B | 124.6 (69.02) | $6.074 \times 10^{-17}$ |
| HOMA2-IR | 1.76 (1.82) | $3.7 \times 10^{-2}$ |
| HOMA2-%S | 88.7 (70.94) | $1.4 \times 10^{-1}$ |
| eGFR (mL/min/1.73m<sup>2</sup>) | 109.9 (14.8) | $4.8 \times 10^{-2}$ |

The clustering analysis included 2,687 (97%) T2D and 68 (95%) T1D individuals which had all variables required for clustering. Using k-means clustering, we categorized T2D into four subtypes (Fig. 1). Consistent with previous studies, SIDD exhibited the highest HbA1C levels. Both MOD and SIRD had the highest BMI among the groups. Notably, MOD individuals were diagnosed at an older age compared to other subtypes, whereas SAID individuals had the earliest diagnosis. SIRD individuals demonstrated the highest values for insulin resistance (HOMA2-IR) and beta-cell function (HOMA2-%B). The distribution of T2D subtypes within the cohort showed that MOD was the most prevalent subtype at 39%, followed by MARD at 36%, SIDD at 18%, and SIRD at 4%.

**Figure 1:**
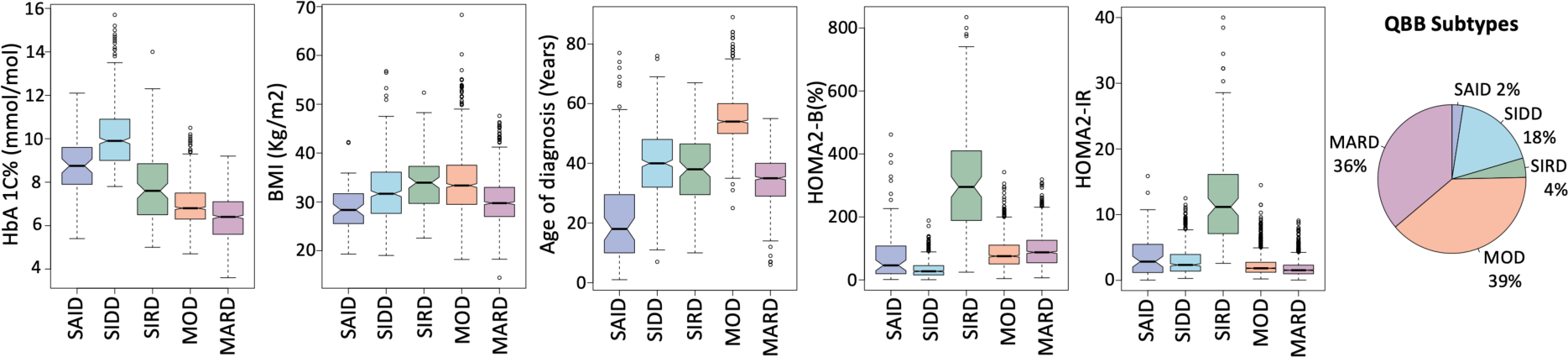
Distribution of Type 2 diabetes subtypes in the QBB derived by k-means clustering. This figure illustrates the subtypes of diabetes among 2,687 clustered individuals with T2D and 68 with SAID from the 13,808 participants Qatar Biobank (QBB) cohort. The x-axis categorizes the subtypes, and the y-axis displays corresponding values of HbA1C, BMI, age of diagnosis, and HOMA2 levels (HOMA2-%B and HOMA2-IR). In this cohort, SIDD presents the highest HbA1C. MOD and SIRD subtypes show the highest BMI. SAID individuals have the youngest age of diagnosis, whereas MOD individuals have the oldest. SIRD shows the highest levels of HOMA2-%B and HOMA2-IR. QBB: Qatar Biobank; SAID: Severe autoimmune diabetes; SIDD: Severe insulin-deficient diabetes; SIRD: Severe insulin-resistant diabetes; MOD: Mild obesity-related diabetes; MARD: Mild age-related diabetes.

### Assessing the cluster reproducibility by applying coordinates from ANDIS cohort

We applied the ANDIS cohort coordinates to the QBB individuals to assess the reproducibility of our data-driven clusters (Fig. S1a). Contrary to previous reports (4) that identified MOD as having the lowest age of diagnosis, our use of ANDIS coordinates revealed MARD as the predominant subtype, accounting for 44%. To visualize the shifts in cluster assignment within the QBB when employing ANDIS coordinates compared to original QBB clusters, a sankey plot was used (Fig. S2a). Notably, SIDD subtype demonstrated the most stability in cluster membership. While only 4% of individuals were categorized as SIRD using QBB coordinates, this figure rose to 11% with ANDIS coordinates, as numerous individuals initially classified as MOD under QBB were reassigned to SIRD using ANDIS. Over half of the initial clusters for MOD and MARD experienced changes in ANDIS coordinates. The significant shift underscores the advantages of employing population-specific derived cluster centers over using coordinates from disparate population studies.

### Assessing the Impact of Using Actual Age versus Age of Diagnosis in Clustering

To evaluate the impact of replacing the age of diagnosis with actual age on clustering, we analyzed a subset of 1,772 individuals (65% of our sample) with complete data for five variables: HbA1C, Age of diagnosis, BMI, HOMA2-%B, and HOMA2-IR. Clustering was first performed using the recorded age of diagnosis, followed by a second analysis in which this variable was substituted with the participants’ actual age. The results revealed notable differences in clustering outcomes depending on the age metric used. When the age of diagnosis was applied, MOD was identified as the predominant subtype (Fig. S1b). In contrast, using actual age shifted the classification, making MARD the dominant subtype (Fig. S1c). A sankey plot visualized the retention and changes in cluster assignments when switching from the age of diagnosis to the actual age (Fig. S2b). The analysis showed that SIDD and SIRD were more stable, maintaining consistent memberships, while MOD and MARD exhibited greater variability, retaining only about 50% of their original clusters. This finding highlights the sensitivity of MOD and MARD classifications to the choice of age metric. The results emphasize the significant impact of using actual age on clustering outcomes for these subtypes, highlighting the importance of carefully selecting the age variable in diabetes subtype analysis.

### Identifying Sex Differences in T2D Subtypes within the QBB Cohort

To identify sex differences within the QBB cohort, we examined variations in five variables (HbA1C, BMI, Age of diagnosis, HOMA2-%B, and HOMA2-IR). We also compared individuals having T2D with a control group of normal individuals, segmented by sex, to ensure the identified differences were unique to T2D (Fig. 2). Significant sex disparities emerged predominantly in the MARD subtype, where males exhibited higher HbA1C levels than females. Across all subtypes (SIDD, SIRD, MOD, MARD), females consistently showed higher BMI values than their male counterparts. The analysis of age metrics revealed that MOD individuals were generally older at diagnosis compared to other subtypes. Specifically, the average age at diagnosis for MOD males was 55.7, increasing to 59.5 years when considering actual age. For MOD females, these figures were 54.5 and 57.8 years, respectively. In contrast, MARD males had an average age of diagnosis of 36.4 years, which rose to 46.1 years for actual age, while MARD females were diagnosed at an average age of 33.7 years and had an actual age of 43.4 years. In terms of beta-cell function, MARD females exhibited significantly higher HOMA2-%B levels than MARD males, highlighting a notable sex difference within this subtype. However, no significant differences were observed in HOMA2-IR levels between the sexes (Table S1). This analysis underscores the importance of considering sex differences in the clinical evaluation and management of diabetes, as these variables influence disease presentation and progression significantly within different T2D subtypes.

**Figure 2:**
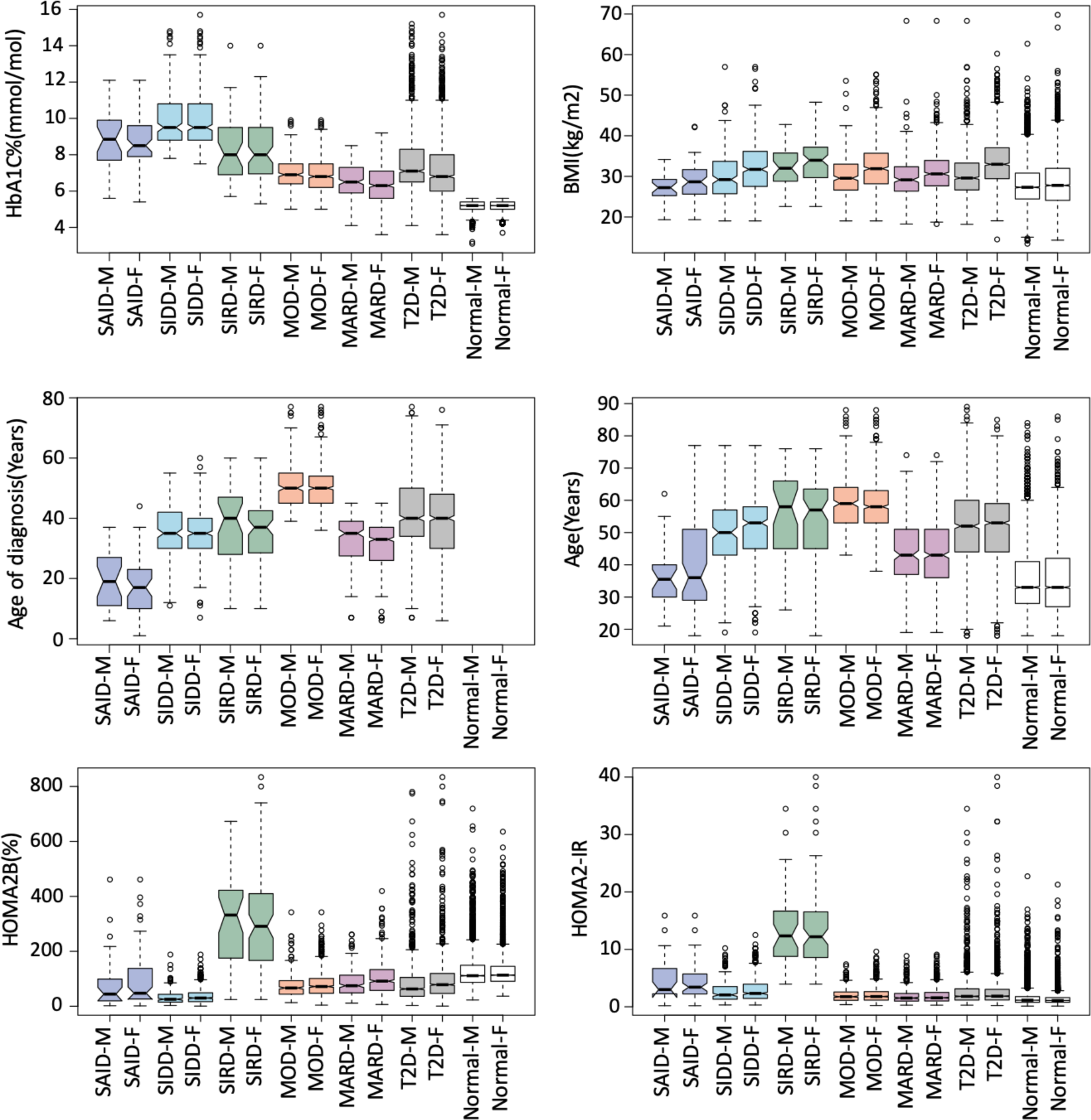
Distribution of Key Variables by Sex Across Diabetes Subtypes in the QBB Cohort. This figure displays the distribution of HbA1C, BMI, age of diagnosis, age, and HOMA2 levels (HOMA2-%B and HOMA2-IR) across T1D, T2D subtypes, and normal, categorized by sex. The x-axis categorizes the subtypes (M: male, F: female), and the y-axis shows the corresponding values for each metric. HbA1C levels are relatively consistent between sexes across subtypes, with SIDD recording the highest levels. Females across all four subtypes demonstrate higher BMI than their male counterparts. The age of diagnosis is comparatively lower for both males and females in MARD, whereas MOD individuals are diagnosed at an older age, regardless of sex. Additionally, MOD individuals tend to be older, and MARD individuals younger, within the QBB cohort. Notably, SIRD exhibits significantly elevated HOMA2-%B and HOMA2-IR levels for both males and females.

### Insulin Treatment Across T2D Subtypes in the QBB Cohort

In the QBB cohort, both SIDD and SIRD subtypes displayed significantly higher HbA1C levels compared to those with MOD and MARD. Consequently, it was anticipated that SIDD and SIRD would require more frequent insulin treatment to manage their elevated HbA1C levels. According to the responses from the QBB questionnaire, a high percentage of insulin prescriptions were indeed observed in these groups. Specifically, insulin was administered to nearly all SAID individuals at a rate of 97%. For the T2D subtypes, the distribution of insulin treatment was as follows: 41.4% for SIDD, 62.9% for SIRD, 12.8% for MOD, and 14.4% for MARD. This pattern underscores the need for targeted insulin therapies, particularly in subtypes where hyperglycemia is more pronounced.

### Aligning of MODY Individuals with other T2D subtypes

Our previous study (14) identified 24 individuals who were initially misclassified as having T2D, but were later confirmed to have maturity-onset diabetes of the young (MODY) through genetic analysis. In our current analysis, we extracted these individuals and analyzed their data across five parameters: HbA1C, BMI, Age of diagnosis, HOMA2-%B, and HOMA2-IR as shown in Fig. 3a. The analysis revealed that MODY individuals typically exhibit lower levels of HbA1C, BMI, HOMA2-%B, and HOMA2-IR, which more closely resemble the characteristics of the MARD subtype. Furthermore, when clustering MODY among the T2D individuals, the majority of MODY cases fell under the MARD category, with a smaller number aligning with MOD and SIDD subtypes as depicted in Fig. 3b. This approach helps delineate the distinct metabolic profiles of MODY within the broader context of T2D subtypes.

**Figure 3:**
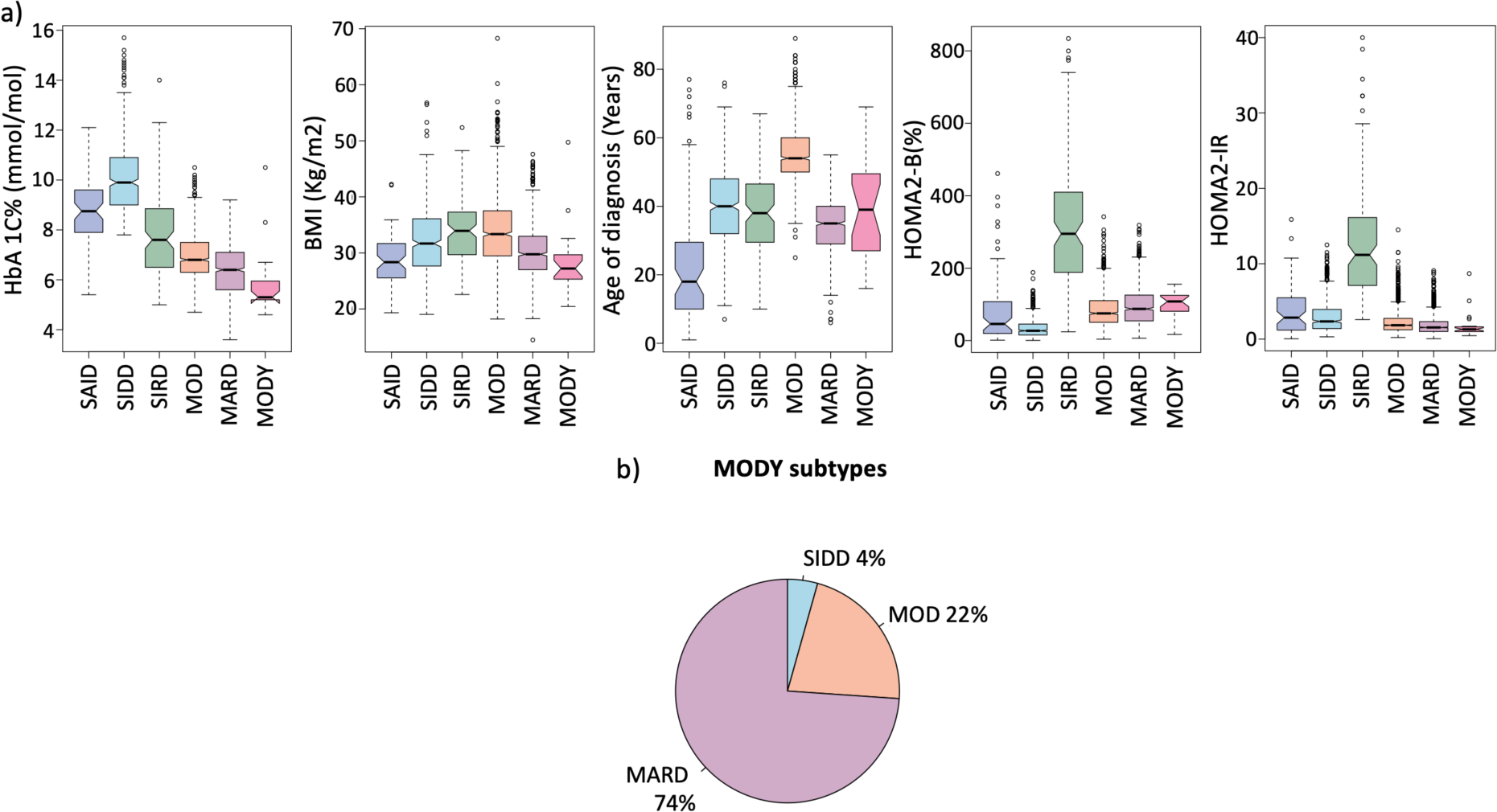
Comparative Analysis of MODY individuals and T2D subtypes in the QBB cohort. a) An assessment was conducted on the 24 individuals diagnosed with MODY, evaluating them across the five variables. This comparison against the four T2D subtypes revealed that MODY individuals generally exhibited the lowest levels of HbA1C, BMI, and HOMA2-IR. b) When the cluster coordinates of the QBB cohort were applied to the MODY group, the majority of MODY cases were categorized under the MARD subtype with 17 out of 24 individuals (approximately 74%) falling into this category. A smaller number of individuals were classified as having MOD with 5 individuals (22%), and only one was categorized as SIDD representing 4% of the MODY group.

### Assessment of Chronic Kidney Disease Prevalence in the QBB Cohort

We analyzed the prevalence and stage of Chronic Kidney Disease (CKD) across the QBB cohort. CKD was categorized into five stages based on estimated Glomerular Filtration Rate (eGFR): stage 1 indicating normal kidney function (eGFR > 90 ml/min/1.73 m^2^), stage 2 for mild loss (eGFR 89 to 60 ml/min/1.73 m^2^), stage 3A for moderate loss (eGFR 59 to 45 ml/min/1.73 m^2^), stage 3B for more significant moderate loss (eGFR 44 to 30 ml/min/1.73 m^2^), and stage 4 for severe loss (eGFR 29 to 15 ml/min/1.73 m^2^). The analysis revealed that a significant number of T2D individuals progressed to various stages of CKD, with most T2D cases falling within stages 3A or higher (Figure 4a). In contrast, individuals without diabetes (Normal) predominantly remained within the normal or mild loss categories. Pre-diabetic individuals primarily exhibited stage 2 CKD, with a minority progressing to more severe stages.

**Figure 4:**
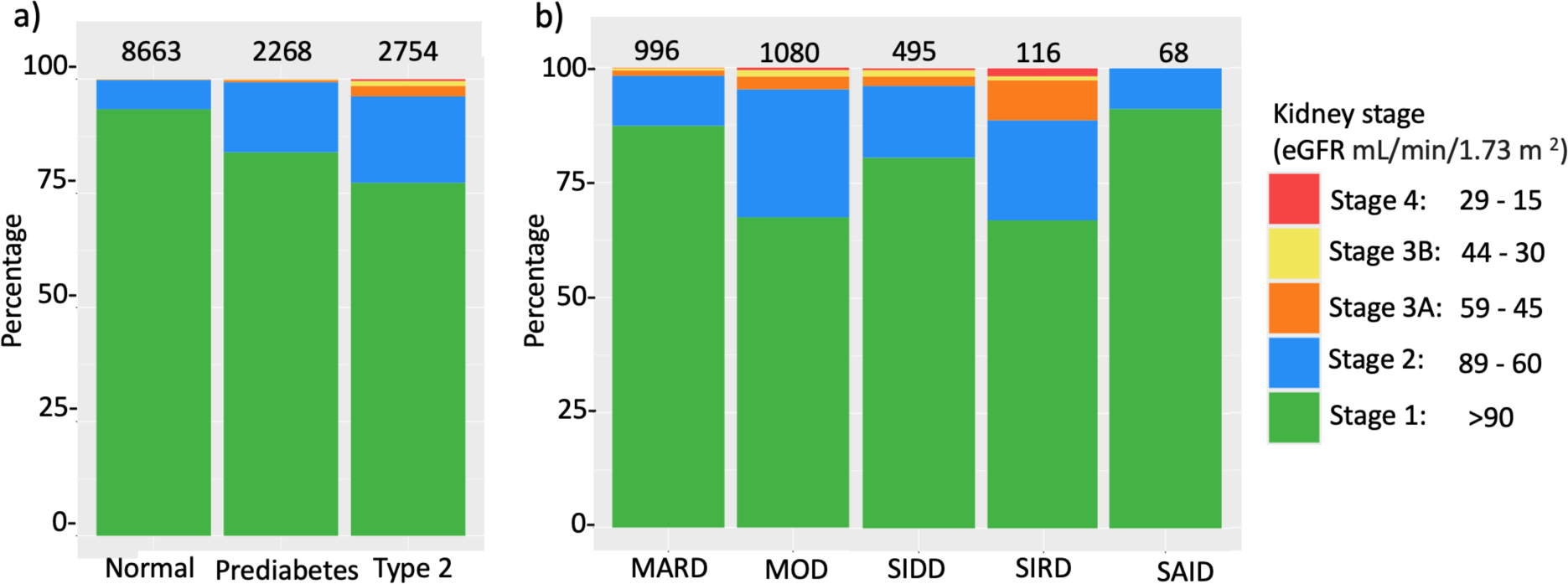
Staging of CKD Across Diabetes and the Subtypes. The x-axis lists the groups, and the y-axis indicates the percentage while the total number of individuals listed above. a) This panel illustrates the distribution of kidney function among individuals categorized as Normal, Prediabetes, and all T2D subtypes combined. The majority of normal individuals had minimal kidney function loss. Prediabetes individuals showed a higher prevalence of kidney damage, while T2D individuals predominantly exhibit significant kidney function impairment. b) This panel details the kidney function across the four subtypes (SIDD, SIRD, MOD, MARD) and SAID. The prevalence of kidney damage was highest in SIRD, followed by MOD, SIDD, and least in MARD. Whereas SAID individuals had normal and mild kidney loss. Color codes for CKD Stages: Stage 1 (green); Stage 2 (blue); Stage 3A (orange); Stage 3B (yellow); Stage 4 (red).

This finding underscores that CKD occurrences are notably higher among individuals with T2D compared to those without diabetes. Further stratification assessed CKD risks within each T2D subtype (Figure 4b). The SIRD subtype showed the highest incidence of stage 3 or higher CKD, whereas a smaller proportion of those with SIDD or MOD were affected. The MARD group displayed the lowest incidence of CKD, and individuals with SAID primarily exhibited mainly normal kidney function and a small proportion with stage 2 CKD. These findings provide critical insights into the differential risks of kidney disease across diabetes subtypes, indicating a need for targeted clinical monitoring and intervention strategies.

We checked for significant differences in the occurrence of CKD among various T2D subtype pairs using Fisher’s exact test (Table 2). Individuals with kidney function stages 1 or 2 were considered healthy, while those in stages 3A, 3B, or 4 were considered diseased. The analysis revealed that SIRD subtype had a significantly higher incidence of CKD compared to SIDD (odds ratio 2.9; p-value 3.8×10^-3^), MOD (odds ratio 2.4; p-value 7.1×10^-3^), and MARD (odds ratio 6.2; p-value 3.36 × 10^-6^). SIDD had a higher incidence compared to MARD (odds ratio 2.1; p-value 2.1×10^-2^), while MOD had a higher incidence compared to SIDD, but not significant (odds ratio 1.2; p-value 5.1×10^-1^). lastly, MOD had a higher incidence compared to MARD (odds ratio 2.6; p-value 1.9×10^-4^).

**Table 2:** Comparison of kidney function between different T2D subtypes: “Healthy,” representing individuals with kidney function in Stages 1 and 2, and “Diseased,” including those with kidney function classified as Stages 3A, 3B, and 4. Results include the p-value from Fisher’s Exact Test alongside the odds ratio and 95% confidence intervals (CI).

| Subtypes |  | P-value | Odds ratio (95% CI) |
| --- | --- | --- | --- |
| SIRD | SIDD | $3.8 \times 10^{-3}$ | 3.17 (1.39 - 7.03) |
| SIRD | MOD | $7.1 \times 10^{-3}$ | 2.56 (1.23 - 4.9) |
| SIRD | MARD | $3.36 \times 10^{-6}$ | 6.91 (3.01 - 15.3) |
| SIDD | MARD | $2.1 \times 10^{-2}$ | 2.17 (1.06 - 4.43) |
| MOD | SIDD | $5.1 \times 10^{-1}$ | 1.24 (0.71 - 2.25) |
| MOD | MARD | $1.9 \times 10^{-4}$ | 2.69 (1.53 - 4.94) |

We further assessed CKD risk in MODY individuals, as detailed in Table S2. Since the majority of MODY individuals clustered into MARD we would expect MODY individuals to have a lower risk of CKD. The findings highlighted that the majority of MODY clustered into MARD maintained normal kidney function (stage 1), in contrast to those in SIDD (stage 3B) or MOD (stages 1, 2, or 4) which had a higher risk of CKD.

## Discussion

In the QBB cohort, MARD individuals were younger than the other subtypes, suggesting that MARD is associated with a healthier, rather than older, demographic. The analysis shows that data-derived centers outperform ANDIS coordinates for classifying T2D in QBB. We expect errors in the reported age of diagnosis mainly influenced the classification of MOD and MARD but did not impact SIDD and SIRD. Despite higher BMIs in females compared to males within the QBB, this did not translate to increased disease severity, as indicated by comparable HbA1C, HOMA2-%B, and HOMA2-IR levels across sexes. We recommend employing clustering methods in populations lacking genetic testing to better identify misclassified MODY individuals within the MARD group. Additionally, CKD incidence was highest in SIRD, followed by SIDD, MOD, and was lowest in MARD.

### Population-specific features of QBB T2D subtypes

In the QBB cohort, the MOD subtype exhibited significantly lower HbA1C levels compared to SIRD subtype, a finding that diverges from results observed in other populations (4, 15). Previous studies (16) have noted a frequent family history of diabetes in MOD but not in MARD. However, a high prevalence of diabetes family history was reported across all four subtypes in the QBB - 87% in SIDD, 88% in SIRD, 81% in MOD, and 90% in MARD, which may be attributed to the high rates of consanguinity in the Qatari population (17).

Several studies (4, 13, 18–20) have shown that MARD typically presents with a later age of diagnosis, yet this trend was not observed in the QBB cohort (Fig. 1). The age of diagnosis for the QBB cohort and the Korean population (13) was lower compared to the ANDIS cohort (4). Additionally, in the QBB cohort, MOD individuals were older than MARD individuals (Fig. 2), suggesting that in the QBB, those classified as MOD are older T2D individuals diagnosed at a later age than those in MARD. This finding is unique to the QBB population and has not been documented before.

Since MOD and MARD are primarily distinguished by their BMI levels – high for MOD and low BMI for MARD - we further analyzed beta cell function using HOMA2-%B. We found that MARD exhibited significantly higher HOMA2-%B compared to MOD (Fig. 1; Fig. 2). However, when using ANDIS coordinates, MOD showed higher HOMA2-%B compared to MARD (Fig. S1a), which may be attributed to the reclassification of some individuals from MARD to MOD (Fig. S2a). In a study of the Indian population, utilizing ANDIS coordinates led to a higher classification of individuals into MOD and fewer into SIDD and MARD, in contrast to classifications using data-derived centroids (21). Therefore, using data-derived centers is recommended for more accurate T2D classification.

### Phenotypic differences between QBB T2D males and females

The majority of T2D males in the QBB cohort had significantly lower BMI than T2D females (Fig. 2; Table S1). Although females had significantly higher BMI, they did not have significantly severe HbA1C levels or HOMA2-IR compared to males. The prevalence of obesity is more common in females than males globally (22). Another notable sex difference is observed in the MARD subtype: females had significantly lower HbA1C levels compared to males, while the opposite was true for their HOMA2-%B levels (Fig. 2; Table S1). This indicates that MARD females had better beta-cell function, enabling them to maintain better control of their HbA1C levels compared to MARD males (Fig. 2).

A similar sex difference in BMI was observed in the Emirati population, where T2D females were more obese than males as well (23). However, to the best of our knowledge, this is the first report of a significant BMI difference across the four subtypes by sex, highlighting the potential need for different personalized treatments based on sex.

### Incidence of Chronic Kidney Disease in T2D subtypes

An increased risk of renal and cardiovascular events is typically observed in SIDD, SIRD, and MOD subtypes, but not in MARD (12). Contrarily, in the Korean population, higher CKD events were noted in the SIRD, SIDD, and MARD subtypes, with MOD showing lower incidence (13). The SIDD and SIRD subtypes had higher events of CVD compared to MOD, which had lower events of CVD (24). Additionally, retinopathy and neuropathy were more prevalent in SIDD, whereas diabetic kidney and fatty liver disease affected individuals in the SIRD group (4).

In the QBB cohort, due to limited data on cardiovascular risk, our analysis primarily focused on assessing the risk of CKD events (Fig. 4). CKD was more prevalent among T2D individuals compared to the other categories (Fig. 4a), with the majority of pre-diabetics experiencing mild loss (stage 2). Further analysis of CKD incidence by subtype (Fig. 4b) revealed the highest rates in SIRD individuals, followed by SIDD and MOD (Table 2). These findings are consistent with previous reports (4, 12) yet contrast with data from the Korean population, which showed a higher risk of CKD in MARD than MOD (13).

### Potential treatment approaches for T2D subtypes

Among the T2D subtypes SIDD and SIRD exhibit severe disease progression. SIRD individuals may particularly benefit from laparoscopic sleeve gastrectomy (LSG). Studies such as (25) have shown that individuals with less than eight years of T2D duration who underwent LSG achieved complete diabetes remission along with significant reductions in weight, dyslipidemia, and hypertension. Furthermore, (26) reported that SIRD individuals experienced substantial improvement in renal function and higher rates of diabetes remission following bariatric surgery. In terms of pharmacological treatment, SIRD individuals have shown positive responses to thiazolidinedione therapy, which effectively lowers HbA1C levels (27). Additionally, SIRD individuals are more susceptible to non-alcoholic fatty liver disease (NAFLD), but treatments such as pioglitazone, SGLT2 inhibitors (SGLT2i), and GLP-1 receptor agonists (GLP-1RA) have proven effective in halting the progression of NAFLD in this group (28).

Recent studies have shown that GLP1-RA treatment results in a stronger glycemic response in females than in males, according to both observational and clinical trials (29). This indicates that females with the SIDD and SIRD subtype may experience more favorable responses to GLP1-RA than their male counterparts. However, additional research is necessary to explore these effects specifically at the subtype levels, rather than for T2D as a whole. Moreover, it is crucial to conduct further studies to monitor inflammatory markers in SIRD individuals, who have shown elevated levels of IL-6, EN-RAGE, and CASP-8 have been reported in this group (30). Investigating potential differences in these inflammatory markers between sexes could yield a deeper understanding of the optimal treatment and management for SIRD.

In the QBB cohort, MARD individuals displayed the lowest incidence of CKD (Fig. 4b; Table 2). These individuals also had lower HbA1C compared to other subtypes (Fig. 1) and demonstrated the highest beta-cell function (HOMA2-%B) (Fig. 2). Research has indicated that the MARD subtype experiences a reduction in fasting glucose following metformin treatment (31). Consequently, MARD individuals in the QBB may particularly benefit from metformin, a first-line medication for diabetes.

Insulin treatment is commonly prescribed for SIDD (41%) and SIRD (62%) individuals in the QBB cohort. However, insulin is associated with cardiovascular risks due to its potential to increase weight, induce hypoglycemia, and cause hyperinsulinemia. In contrast, other T2D medications have been shown to reduce both mortality and cardiovascular events, presenting them as better alternatives to insulin (32). Moreover, T2D individuals with high insulin resistance who are treated with insulin have a heightened risk of diabetic kidney disease and cardiovascular events (33). Additionally, those taking both insulin and other antidiabetic drugs exhibit a higher cardiovascular risk ratio compared to those on antidiabetic drugs alone (34). Therefore, it is advisable to personalize treatment for SIRD individuals, prioritizing medications such as SGLT2 inhibitors and GLP-1RA over insulin. Although SIDD individuals typically have lower BMI compared to SIRD and MOD (Fig. 1), clinicians should closely monitor these individuals for weight gain and evaluate if the benefits of insulin in reducing HbA1C levels outweigh the potential risks.

### Clustering of MODY individuals within the T2D Framework

Our analysis showed that the majority of the 24 MODY individuals in the QBB (14) predominantly clustered into MARD (Fig. 3b). We hypothesize that MODY patients, likely misclassified within the clustering scheme, tend to be grouped into MARD. It has been observed that MARD subtype responds well to sulfonylurea therapy (27). Additionally, individuals with HNF1A and HNF4A mutations show high sensitivity to sulfonylurea treatment (35, 36), and about 45% of the 24 MODY cases had these mutations (14). The tendency for MODY individuals to cluster into MARD could help explain why this subtype derives significant benefits from sulfonylurea therapy, as these individuals may be misclassified MODY patients rather than typical T2D. In populations lacking genetic testing, identifying MODY individuals among T2D patients who cluster into the MARD subtype and respond well to sulfonylurea therapy might be feasible.

In the QBB cohort, while some MODY individuals developed CKD, the majority retained normal kidney function (Table S2). Recent research, including a study on the Indian population (37), has noted a significant occurrence of nephropathy and retinopathy in MODY individuals. This may explain the CKD risk observed among MODY individuals in the QBB cohort (Table S2). However, a more comprehensive understanding of the CKD risk requires additional research, especially with a larger cohort of MODY individuals.

### Study Limitations

Our study has certain limitations, including the absence of risk assessments for NAFLD and CVD for the subtypes and the lack of GADA measurements to confirm SAID individuals. However, it is reassuring that none of the SAID individuals developed CKD, suggesting that the classification is not biased, as T1D does not have a high risk of CKD.

## Conclusion

In our comprehensive study of the Qatari population within the Qatar Biobank, we successfully classified 2,687 individuals with T2D into four distinct subtypes. Our findings illuminate several unique characteristics of these subtypes, thereby enriching the framework for personalized diabetes management in this Middle Eastern population. Notably, our study underscored significant sex-based differences in diabetes expression. Females, across all subtypes, exhibited higher BMI levels than their male counterparts, while MARD females demonstrated higher beta-cell function and lower HbA1C levels compared to MARD males. These sex-specific differences emphasize the need for tailored diabetes treatment strategies that consider both biological and sex-specific factors.

The clustering approach also revealed that many MODY individuals aligned with the MARD subtype, suggesting that misclassified MODY cases within MARD may account for the observed sensitivity to sulfonylurea therapy. This finding highlights the potential role of genetic testing in accurately identifying MODY cases within the T2D framework, which could improve treatment efficacy.

Our CKD analysis showed that SIRD, SIDD, and MOD subtypes face higher CKD risks compared to MARD, with SIRD exhibiting the highest incidence and most severe stages. These findings support the need for intensive monitoring and tailored therapeutic strategies, particularly for SIRD individuals, who may benefit more from treatments like bariatric surgery and medications such as SGLT2 inhibitors and GLP-1 receptor agonists over insulin. For SIDD individuals, personalized insulin management remains crucial to balance glycemic control and minimize associated risks.

Overall, this study supports the use of T2D subtyping to advance personalized treatment strategies, contributing valuable insights to enhance diabetes management in Middle Eastern populations. By refining subtype identification and considering population-specific factors, healthcare providers can offer more effective, individualized interventions that improve patient outcomes and quality of life.

## Methods

### Study Cohort

The QBB cohort is a population-based study in Qatar, that recruited Qatari and long-term residents (>= 15 years living in Qatar) aged 18 years and older (38). Extensive baseline socio-demographic data, clinical and behavioral phenotypic data, and serum concentrations of HbA1c, triglycerides, glucose, C-peptide, creatinine, total cholesterol, LDL-C, and HDL-C, and multiple other clinical biochemistry parameters (39) have been measured at the central laboratory of Hamad Medical Corporation (HMC), accredited by the College of American Pathologists. All participants provided informed consent, and the study received approval from the QBB Institutional Review Board (Protocol no. E-2019-QF-QBB-RES-ACC-0179-0104). No compensation was given to the participants. At the time of this study, QBB data were available for 13,808 Qatari participants. T2D classification was based on self-reported physician diagnosis, use of diabetic medications, HbA1C level > 6.5%, or random glucose levels > 200 mg/dL. Individuals with T1D (SAID) were identified based on insulin treatment and C-peptide levels < 0.5 nmol/L (11).

### Definition of T2D and controls

Subjects were defined as controls if all the following four conditions were satisfied: first, no self-reported physician diagnosis of diabetes; second, no self-reported treatment with any diabetes-specific medication; third, HbA1c < 5.7%; and fourth, random glucose level <200 mg/dL. T2D was defined if any one of the following four conditions was satisfied: first, having a physician diagnosis of diabetes based on the questionnaire (86% of all participants), second, being treated for diabetes based on the QBB questionnaire (62%), third, having an HbA1c > 6.5% (63%), or fourth, having random glucose >200 mg/dL (17%). Based on this definition, X% of individuals were defined as having T2D. Individuals with HbA1c between 5.7% and 6.4% (N = 1845) were excluded. Ahlqvist et al. used glutamic acid decarboxylase antibodies (GADA) to define an additional subtype of SAID. As GADA measurements were not available in QBB, individuals with self-reported type 1 diabetes (T1D) or C-peptide concentrations below 0.5 nmol/L and on insulin treatment were classified as SAID (N = 71).

### Cluster Analysis

K-means clustering was performed using HbA1C, BMI, age of diagnosis, HOMA2-%B, and HOMA2-IR setting the number of clusters (k) to 4 to derive the four subtypes of T2D. For individuals missing the age of diagnosis (35%), their actual age was used as a substitute. HOMA2-%B and HOMA2-IR were calculated for all T2D individuals based on C-peptide levels using the HOMA2 calculator (University of Oxford, Oxford, UK) (40). Patients defined as SAID were excluded from the clustering and assigned to their own subtype, and clustering was carried out on the patients with T2D only. Clustering analysis was carried out on standardized values centered at a mean of 0 and a standard deviation of 1. The optimal number of clusters was determined using the k-means function in the “cluster” and “factoextra” libraries in R (version 4.3.1). we determined the optimal number of clusters to be k = 4. This finding was consistent with that observed by Ahlqvist et al. The cluster variables in QBB followed a similar trend to the ANDIS cohort. Therefore, we assigned cluster labels based on the five clinical variable averages characteristic of each T2D subtype following Ahlqvist et al.

### Sensitivity Analyses

We conducted several sensitivity analyses to assess the robustness of our clustering results. First, we evaluated the impact of using the published ANDIS coordinates (4) compared to our data-derived coordinates for identifying T2D subtypes in the QBB cohort. Changes in cluster assignments were visualized using a Sankey plot to track the flow of cluster membership between QBB and ANDIS coordinates. Second, we examined the effect of different age variables on the clustering outcomes by performing clustering based on the reported age of diagnosis and then repeating the analysis using the actual age of the participants. Both analyses employed the same clustering parameters that were used in the initial analysis. Third, we investigated potential sex differences among the T2D subtypes by splitting the dataset into male and female groups. We compared five variables (HbA1C, BMI, Age of diagnosis, HOMA2-%B, HOMA2-IR) across subtypes, using t-tests to identify significant differences between sexes for each variable.

### Analysis of Insulin Use, MODY Classification, and CKD Risk Across QBB Subtypes

First, we used data from the QBB questionnaire to identify individuals prescribed insulin treatment, enabling a comparison across the subgroups. Second, we compared MODY individuals as identified in (14) with T2D individuals using five variables (HbA1C, BMI, age of diagnosis, HOMA2-%B, and HOMA2-IR). These MODY individuals were then clustered alongside T2D individuals to determine within the subtypes. Third, the incidence of chronic kidney disease (CKD) was assessed by calculating the estimated glomerular filtration rate (eGFR) based on creatinine levels, sex, and age (41, 42). Fisher’s exact test was applied to identify which T2D subtype had a higher risk of CKD.

## DATA AVAILABILITY STATEMENT

The QBB data are available under restricted access for the informed consent given by the study participants does not cover posting of participant-level phenotype data in public databases, access can be obtained in the form of an MS SQL Server 2008 R2 database, upon request from QBB (https://www.qatarbiobank.org.qa/research/how-to-apply)

## ACKNOWLEDGEMENTS

We are grateful to all study participants of Qatar Biobank for their invaluable contributions to this study. This work is supported by a cluster grant from Qatar Research Development and Innovation Council (QRDI) awarded to O.M.E.A, K.S. and A.B.A-S. (NPRP11C-0115-180010). K.S. is supported by the Biomedical Research Program at Weill Cornell Medicine in Qatar, a program funded by the Qatar Foundation. O.M.E.A. is supported by the College of Health and Life science (CHLS) of Hamad Bin Khalifa University (HBKU). K.S. is also supported by the Qatar Research Development and Innovation Council (QRDI) grant ARG01-0420-230007. N.M.A. is supported by the Qatar Genome Program (QGP) and CHLS of HBKU Scholarship.

## AUTHOR CONTRIBUTION STATEMENT

N.M.A: Data curation, Formal analysis, Investigation, Writing – original draft. S.B.Z: Formal analysis, Investigation, Writing – review, and editing. A.B.A-S: Investigation, Resources. K.S. and O.M.E.A: Conceptualization, Funding acquisition, Project administration, Resources, Investigation, Supervision, Writing – review and editing. All authors have read and agreed to the published version of the manuscript.

## COMPETING INTERESTS STATEMENT

The other authors declare no competing interests.

## Notes

### Competing Interest Statement

The authors have declared no competing interest.

### Funding Statement

This study was funded by Qatar Research Development and Innovation Council (QRDI) awarded to O.M.E.A, K.S. and A.B.A-S. (NPRP11C-0115-180010). K.S. is supported by the Biomedical Research Program at Weill Cornell Medicine in Qatar, a program funded by the Qatar Foundation. O.M.E.A. is supported by the College of Health and Life science (CHLS) of Hamad Bin Khalifa University (HBKU). K.S. is also supported by the Qatar Research Development and Innovation Council (QRDI) grant ARG01-0420-230007. N.M.A. is supported by the Qatar Genome Program (QGP) and CHLS of HBKU Scholarship.

### Author Declarations

Ethics committee/IRB of Qatar BioBank gave ethical approval for this work

