## Supplementary material for "Subtyping of Type 2 Diabetes from a large Middle Eastern Biobank: Implications for Precision Medicine": Fig. S1, Fig. S2, Table S1, Table S2

**List:**

Figure S1: Clustering of QBB individuals under different settings

Figure S2: Changes in cluster membership due to varying clustering metrics.

Table S1: Comparison of cluster variables between males and females across different T2D subtypes.

Table S2: Data values for the 24 MODY individuals in QBB.

**Figures:**

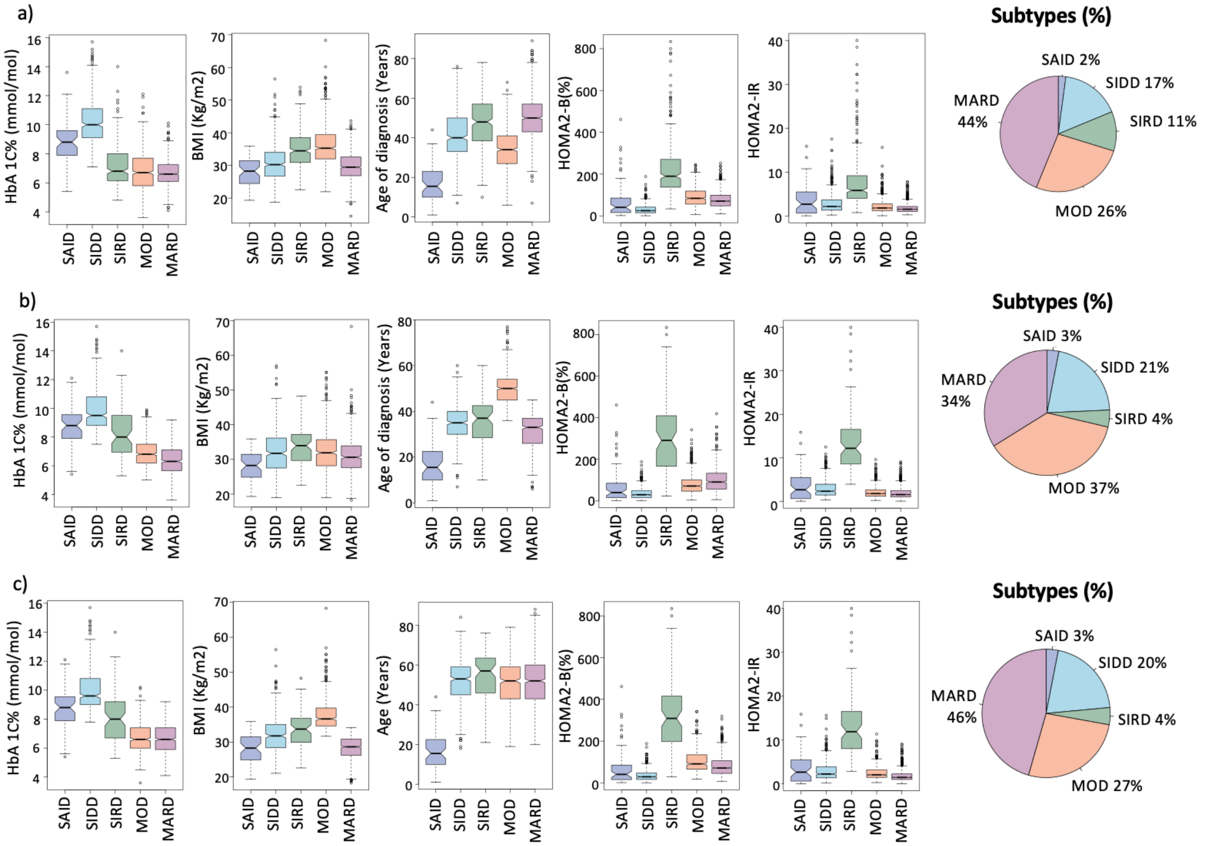


**Figure S1: Clustering of QBB individuals under different settings.** The x-axis lists subtypes, while the y-axis represents values of HbA1C, BMI, age of diagnosis, and HOMA2 levels (HOMA2-%B and HOMA2-IR). a) QBB subtypes were identified using ANDIS-derived coordinates. When using ANDIS coordinates, MOD had a low age of diagnosis, and MARD had a later age of diagnosis. The major subtype in QBB when using ANDIS coordinates was MARD. b) QBB subtypes were identified using the available age of diagnosis for 1,772 individuals. The subtype features followed a similar distribution to the original clustering in QBB. c) QBB subtypes were identified using actual age instead of age of diagnosis for the same 1,772 individuals. After substituting age of diagnosis with age, MOD had the highest BMI and MARD was the largest subtype.

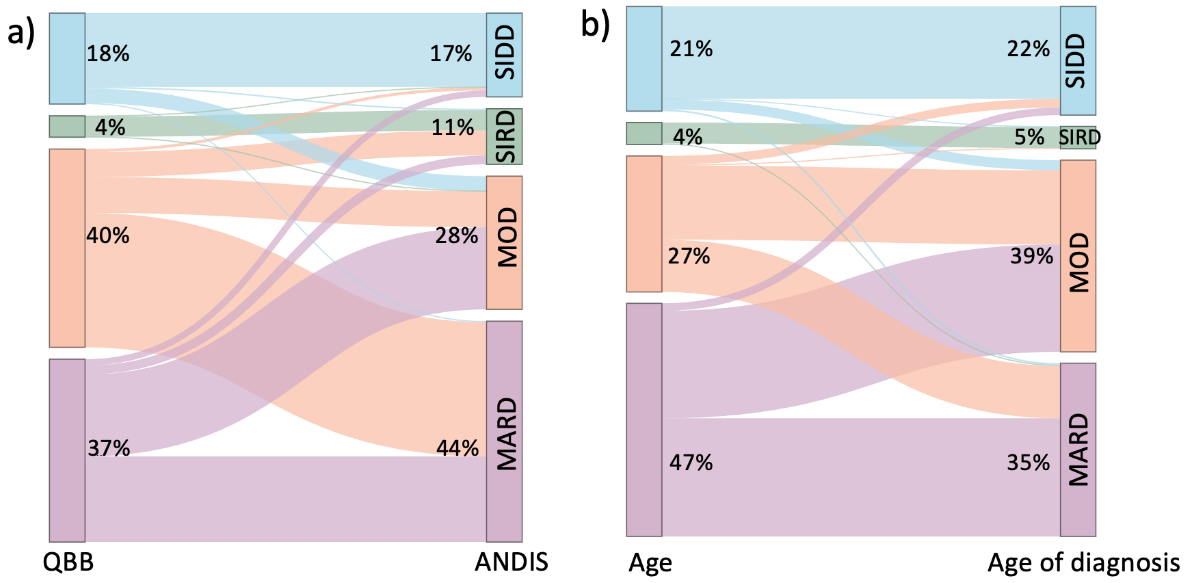


**Figure S2: Changes in cluster membership due to varying clustering metrics.** a) Cluster changes when using QBB-derived centers as opposed to using ANDIS coordinates. b) Cluster changes of T2D individuals when using the actual age instead of age of diagnosis.

**Tables:**

**Table S1: Comparison of cluster variables between males and females across different T2D subtypes.** This table displays p-values calculated using t-tests to compare data points between males and females within each T2D subtype. Significant differences in HbA1C were observed only for MOD and MARD subtypes. Significant differences in BMI were observed between males and females across all four T2D subtypes. For Age of diagnosis, we observed significant differences for MOD and MARD subtypes. Significant differences for HOMA2-%B were observed only for SIDD and MARD subtypes. Significant differences in HOMA2-IR were observed only for SIRD subtype.

| | HbA1 C% | BMI | Age of diagnosis | HOMA2-%B | HOMA2-IR |
|---|---|---|---|---|---|
| SIDD | $9.95\times10^{-1}$ | $3.82\times10^{-13}$ | $8.7\times10^{-1}$ | $7.146\times10^{-3}$ | $1.572\times10^{-2}$ |
| SIRD | $9.994\times10^{-1}$ | $2.288\times10^{-2}$ | $1.2\times10^{-1}$ | $6.113\times10^{-1}$ | $2.727\times10^{-1}$ |
| MOD | $2.432\times10^{-2}$ | $< 2.2 \times 10^{-16}$ | $2.399\times10^{-2}$ | $2.288\times10^{-1}$ | $4.717\times10^{-1}$ |

| | | | | | |
|---|---|---|---|---|---|
| MARD | $6.11 \times 10^{-11}$ | $3.2 \times 10^{-12}$ | $6.85 \times 10^{-7}$ | $2.92 \times 10^{-8}$ | $6.989 \times 10^{-1}$ |

**Table S2: Data values for the 24 MODY individuals in QBB**. The Average Age / Age at diagnosis for MODY individuals was 38 years (± 14 Standard Deviation), BMI, HbA1C, HOMA2 estimates, and subtype of each MODY cluster. The stages of chronic kidney disease (CKD) are listed for all MODY individuals in the QBB cohort. Missing variables for 1 individual are listed as (-). All MODY individuals clustered into MARD had normal kidney function (stage 1). MODY individuals clustered into MOD and SIDD had mild, moderate and severe kidney loss.

| BMI | HbA1 C% | HOMA2-%B | HOMA2-IR | Assigned Subtype | CKD Stage | Gene |
|---|---|---|---|---|---|---|
| 27.21 | 10.5 | 17.1 | 8.695652 | SIDD | 3B | HNF4A |
| 37.56 | 8.3 | 79.8 | 5.076142 | MOD | 4 | HNF1A |
| 30.52 | 6.7 | 82.2 | 2.680965 | MOD | 2 | GCK |
| 32.57 | 5.4 | 127.3 | 1.342282 | MOD | 2 | HNF1A |
| 28.55 | 6.5 | 60.9 | 1.129944 | MARD | 1 | GCK |
| 25.43 | 6.4 | 73.1 | 1.293661 | MARD | 1 | GCK |
| 23.26 | 5.1 | 97.8 | 0.638978 | MARD | 1 | GCK |
| 27.81 | 5.9 | 34.8 | 0.455581 | MARD | 1 | HNF1A |
| 24.63 | 4.6 | 108.1 | 1.204819 | MOD | 1 | HNF1A |
| 49.74 | 5.8 | 116.9 | 1.52439 | MOD | 1 | HNF1A |
| 20.44 | 5.2 | 120.6 | 1.010101 | MARD | 1 | HNF1A |
| 26.03 | 5.6 | 76.3 | 1.373626 | MARD | 1 | HNF1A |
| 26.18 | 5.3 | 121.5 | 1.517451 | MARD | 1 | HNF1A |
| 28.58 | 5.2 | 155.7 | 1.574803 | MARD | 1 | HNF1A |
| 29.06 | 5.1 | 137.8 | 1.709402 | MARD | 1 | HNF1A |
| 23.97 | - | - | - | - | 1 | KLF11 |
| 30.92 | 5.3 | 119.2 | 1.066098 | MARD | 1 | BLK |
| 25.45 | 5.2 | 141.5 | 2.898551 | MARD | 1 | BLK |
| 20.89 | 5.1 | 98.5 | 0.597729 | MARD | 1 | BLK |
| 29.55 | 5.3 | 124 | 0.978474 | MARD | 1 | BLK |
| 22.07 | 5.1 | 149.3 | 1.692047 | MARD | 1 | BLK |
| 25.2 | 5.7 | 86.3 | 0.761035 | MARD | 1 | ABCC8 |
| 26.01 | 6 | 100.5 | 0.717875 | MARD | 1 | ABCC8 |
| 29.79 | 5.3 | 125.8 | 1.234568 | MARD | 1 | ABCC8 |
